# Lipid profiling reveals a deficit of lipids with unsaturated fatty acids in women with Alzheimer’s disease

**DOI:** 10.1101/2025.02.26.25322937

**Authors:** Asger Wretlind, Jin Xu, Wenqiang Chen, Latha Velayudhan, Petroula Proitsi, Cristina Legido-Quigley, the AddNeuroMed Consortium

## Abstract

**Introduction:** Alzheimer’s disease (AD) is a devastating neurological disease that disproportionately affects women. This study aimed to investigate sex-specific single lipids associated with AD.

**Methods:** Plasma samples from 841 participants, comprising 306 individuals with AD, 165 with mild cognitive impairment (MCI), and 370 cognitively healthy controls were curated from the AddNeuroMed cohort. Lipidomics identified 268 single lipids for each sample. We investigated sex-specific associations from lipid modules and single-lipids to AD and probed for causality with mediation analyses.

**Results:** Three modules associated with AD in the female subset and one in the male subset (p < 0.05). In the female participants with AD, lipid families containing highly unsaturated fatty acids were reduced and those containing saturated lipids were increased (q-value < 0.05). The effects of unsaturated phospholipids on AD were not mediated via cholesterol, LDL or ApoB.

**Discussion:** Women with AD have lower unsaturated blood lipid levels compared to controls.

## 1. Introduction

Alzheimer’s disease (AD) is a devastating neurodegenerative disease that affects an increasing number of people worldwide. Women are disproportionately impacted by AD, accounting for approximately two-thirds of all AD cases (1,2). While women’s longer lifespan has been suggested as an explanation of this overrepresentation, the answer appears to be more complex. Research by Matthews *et al.* (3) and Beam *et al.*(4) demonstrate that women exhibit a higher incidence of AD compared to men after age 80, indicating that longevity alone does not account for this gender disparity. Although sex-dependent biological mechanisms related to AD pathogenesis have been investigated (5), the reasons for women’s increased susceptibility to AD remain unclear (6).

One promising avenue for understanding sex differences in AD risk comes from the identification of biomarkers through metabolomics and lipidomics. Recent metabolomic analyses have illustrated how metabolites associated with AD risk are often sex-specific. For example, Varma *et al.* found that lower levels of bile acids were associated with neuroimaging markers of dementia, with pharmacological reduction of the bile acids, cholic acid and chenodeoxycholic acid, led to increasing risk of vascular dementia primarily in men (7). We recently demonstrated that vanillylmandelate, tryptophan betaine, and kynurenate, metabolites closely related to neurotransmission and inflammation, exhibit sex-specific alterations that could improve predictive modeling in women but not in men (8). Arnold *et al.* further revealed sex differences in 15 metabolites related to AD risk, showing that alterations in some metabolites, for instance, valine, glycine and proline, were evident only in sex-stratified analyses (9). These findings suggest that women may experience a greater impact from impaired mitochondrial energy production in AD, which affects fatty acid metabolism and manifests as differences in the lipidome.

Metabolic difference between women and men, especially in lipid metabolism, are well-documented (10,11). Liu *et al.* found that increased levels of small and medium low-density lipoprotein (LDL) were associated with cognitive decline in women but not in men (12), a finding further supported by Zarzar *et al.* showing that small and medium LDL correlated with AD and mild cognitive impairment (MCI) in women, whereas men showed an association between LDL and MCI with larger LDL particles (13). Importantly, the recent Lancet

Commission for Dementia estimated that 45% of AD risk is potentially modifiable, with 7% attributed to LDL levels (14). Hence, lipid metabolism is crucial for brain health, and lipidomic analysis presently provides the most advanced method for examining lipid changes during AD pathogenesis. As lipidomic technologies continue to advance, enabling the identification and quantification of an increasing number of lipid molecules, we are poised to gain deeper insights into the complex lipid alterations associated with AD pathogenesis in both men and women.

In this study the lipidome of 841 participants at three different levels of cognitive health: healthy, MCI and AD were investigated and lipids association to AD were uncovered. Furthermore, we demonstrated that these associations vary by sex and depend on lipid saturation levels.

## 2. Methods

### 2.1 Participants

A post-hoc analysis of data from the AddNeuroMed cohort and the Dementia Case Register cohort (15) was performed. This cohort consisted of 841 participants recruited from six European countries (England, Finland, France, Greece, Italy and Poland). Participants were divided into three groups: 306 were diagnosed with AD according to the criteria of the National Institute of Neurological and Communicative Disorders and Stroke and the Alzheimer’s Disease and Related Disorders Association (NINCDS-ADRDA) and the fourth edition of the Diagnostic and Statistical Manual of Mental Disorders (DSM-IV) (16,17). Additionally, 165 participants were classified as having MCI following evaluation with the Clinical Dementia Rating Scale (CDR) (18) and criteria outlined by Petersen et al. (19). Finally, 370 cognitively healthy participants were recruited as controls, all of whom passed the Mini-Mental State Examination (MMSE) (20) or the Alzheimer’s Disease Assessment Scale - Cognitive Section (ADAS-cog) (21) without signs of cognitive impairment. Notably, individuals with other psychiatric or neurological illness were excluded. Nightingale Health provided measures for LDL, HDL, total cholesterol, total triglycerides and ApoB.

### 2.2 Lipidomics

Blood samples were collected following a recommended two-hour fasting period. Plasma samples were prepared by centrifugation using ethylenediamine tetraacetic acid (EDTA). Lipid extraction and measurement protocols have been previously described in detail (22,23). In brief, lipids were extracted from plasma using methanol to precipitate proteins, followed by a two-phase extraction using methyl tert-butyl ether (MTBE) and water (24). Samples were analyzed using ultra-performance liquid chromatography (UPLC) in tandem with a quadrupole time-of-flight mass spectrometer (QTOF-MS) in both positive and negative ionization modes. A total of 278 lipids were annotated by matching to an in-house library, followed by a manual curation of each chromatogram using Skyline v. 22.1 (25). Lipid peak areas were normalized to exogenously added internal standards, and a final quality control (QC) check was performed to address missing data and outliers. A total number of 268 lipid species passed the QC and were included in the statistical analysis.

### 2.3 Statistics

Data quality was assessed prior to statistics analysis. Variables with more than 20% missing was discarded, while those variables with missing data less than 20% were imputed using k-nearest neighbor (KNN) imputation. Outliers, defined as values outside the 0.01%-99.99% quantile range were winsorized. All lipid data were log10-transformed to achieve a normal distribution. All data preprocessing, statistical analyses, and visualizations were conducted using R v.4.3.0 (26), and the code is available on GitHub: https://github.com/Asger-W/AddNeuroMed-Lipidomics.

Participant characteristics were presented by the three groups of neurological decline status (Control, MCI and AD). The three groups were compared using Welch’s *t*-test for continuous variables and the χ^2^ test for categorical variables using the R package “tableone” (27). Each lipid was regressed against participant age and sampling site, and the residuals were used for weighted correlation network analysis (WCNA), a method originally developed for investigating genetic co-expression that has proven effective for highly correlated data, such as lipidomics (28,29). A signed correlation network was constructed, and highly correlated lipids were clustered into 11 modules using the WCNA package in R (30). Each modulés eigengene, consisting of the summarized data within each module, were regressed against AD, MCI, APOE genotype and sex. Furthermore, data were stratified into female and male subsets for further regression analyses of module eigengenes against AD. All module eigengene regressions were adjusted for covariates not used as predictor variables. For instance, in the regression of AD versus control, adjustment were performed for APOE genotype and sex, as follows: Module_eigengene ∼ AD_status + APOE_genotype + sex.

Modules associated to AD were included for further analysis, focusing on the lipids within these modules. Individual lipids were regressed similarly to the module eigengenes against AD, MCI, APOE genotype, sex, MMSE score, as well as female/male subsets against AD. Analyses were adjusted for potential confounders (AD status, APOE genotype, sex) and p-values were corrected for multiple testing using the False Discovery Rate (FDR). A sensitivity analysis for AD were also carried out to test the impact of each individual confounder, as well as total triglycerides. Differences in lipid levels between individuals with AD and healthy controls were tested separately for female and male participants using Students *t*-test, adjusted for multiple-testing using FDR correction. To evaluate whether the association between lipid molecules and AD is mediated through changes in lipid related and atherosclerosis risk biomarkers (total cholesterol, LDL, HDL, and ApoB), we performed causal mediation analysis. This was implemented using linear regression models in the R package ‘mediation’ (31), with 500 bootstrapped iterations to ensure robust estimates of average direct effects (ADE), total effects and average causal mediated effect (ACME) as described previously (32). Proportion was calculated as total effect divided by the indirect effect.

## 3. Results

### 3.1 Participant characteristics

We investigated circulating lipid molecules among participants at three stages of cognitive decline: healthy control (n = 370), MCI (n = 165), and AD (n = 306). The average age of AD diagnosis was 73.04 years and the average AD duration was 3.74 years. Participants with AD were on average older than their control and MCI counterparts and had a higher proportion of APOE-ε4 carriers (Table 1), consistent with the trends observed in the general AD population (33,34). The results show that, compared to the control group, the AD group had a significantly higher levels of total cholesterol and LDL, though no significant differences were observed in HDL or total triglycerides. Female participants had a higher level of cholesterol, LDL and HDL compared to the male participants, whereas no differences were found in total triglycerides between sexes (Supplementary table 1).

**Table 1.** Participant characteristics. Data are presented as n (%), mean (SD), Groupwise comparisons between the two treatments were tested using a Welch two-sample t test for continuous variables and x^2^ test for categorical variables.

|  | Overall | Ctrl | MCI | AD | Groupwise comparison |
| --- | --- | --- | --- | --- | --- |
| n | 841 | 370 | 165 | 306 |  |
| Age, mean (SD) | 76.24 (6.76) | 75.43 (6.79) | 75.95 (6.68) | 77.37 (6.63) | 0.001 |
| Sex (%) |  |  |  |  | 0.339 |
| Female | 491 (58.4) | 220 (59.5) | 88 (53.3) | 183 (59.8) |  |
| Male | 350 (41.6) | 150 (40.5) | 77 (46.7) | 123 (40.2) |  |
| APOE-ε4 (%) |  |  |  |  | <0.001 |
| Absent | 456 (60.4) | 236 (73.8) | 92 (66.2) | 128 (43.2) |  |
| Heterozygote | 245 (32.5) | 75 (23.4) | 41 (29.5) | 129 (43.6) |  |
| Homozygote | 54 (7.2) | 9 (2.8) | 6 (4.3) | 39 (13.2) |  |
| Total Cholesterol, mean (SD) | 5.40 (1.22) | 5.24 (1.20) | 5.52 (1.31) | 5.52 (1.17) | 0.005 |
| LDL mmol/L, mean (SD) | 2.05 (0.58) | 1.97 (0.57) | 2.11 (0.60) | 2.10 (0.57) | 0.004 |
| HDL mmol/L, mean (SD) | 1.60 (0.38) | 1.59 (0.39) | 1.59 (0.40) | 1.63 (0.37) | 0.369 |
| Total Triglyceride mmol/L, mean (SD) | 1.42 (0.64) | 1.41 (0.63) | 1.48 (0.72) | 1.39 (0.61) | 0.336 |
| ApoB mmol/L, mean (SD) | 0.97 (0.26) | 0.93 (0.25) | 1.00 (0.28) | 0.99 (0.25) | 0.003 |
| MMSE Score, mean (SD) | 25.46 (4.99) | 28.47 (2.67) | 26.80 (2.06) | 20.87 (5.07) | <0.001 |
| Education Years, mean (SD) | 9.95 (4.32) | 11.19 (4.34) | 9.49 (4.05) | 8.86 (4.09) | <0.001 |
| Marital Status (%) |  |  |  |  | <0.001 |
| Divorced | 31 (4.7) | 20 (7.7) | 6 (4.1) | 5 (1.9) |  |
| Married | 394 (59.2) | 157 (60.2) | 93 (63.7) | 144 (55.6) |  |
| Single | 41 (6.2) | 23 (8.8) | 6 (4.1) | 12 (4.6) |  |
| Widowed | 200 (30.0) | 61 (23.4) | 41 (28.1) | 98 (37.8) |  |

### 3.2 Lipid modules associate to AD

A scale-free network of pair-wise Pearson correlations were constructed for the lipids, revealing that lipids within the same lipid family were highly correlated and tended to cluster together in the network (Figure 1.A). Additionally, related lipid families, such as phosphtatidylcholines (PCs) and phosphtatidylethanolamines (PEs), or sphingomyelin (SM) and ceramides (Cer), also clustered together. Modules identification resulted in 12 modules, which was reduced to 11 modules after merging two highly similar modules. The identified modules showed substantial overlap with the lipid families, but these modules also revealed individual lipids that correlated modules of different lipid families (Figure 1.B). Furthermore, the modules can divide clusters of lipid families, for example, triglycerides (TGs) were divided across three TG-dominant modules: M2, M5, and M8.

**Figure 1.**
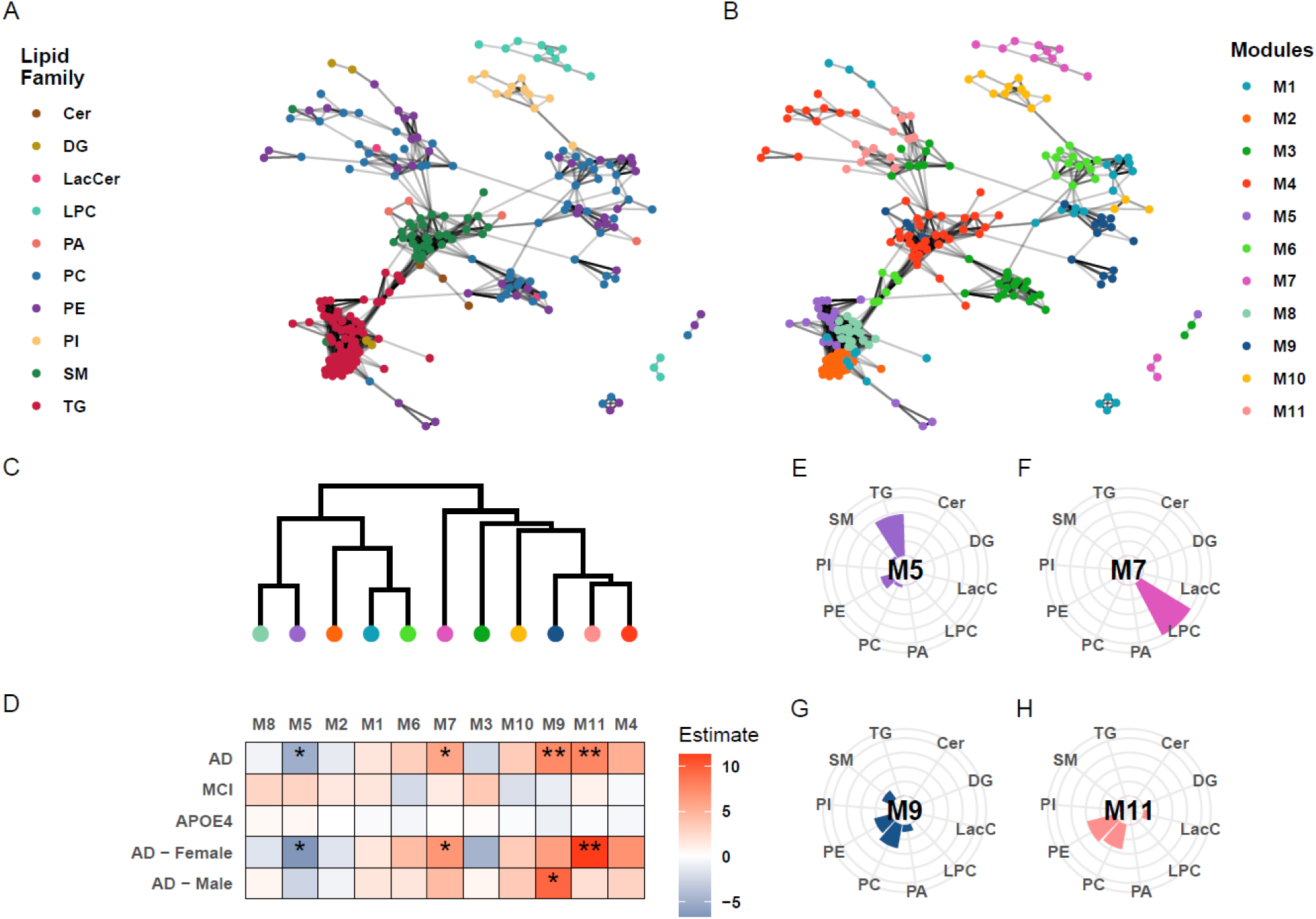
WCNA analysis of lipid species. (A) Correlation network of individual lipid molecules, colored by lipid family. (B) Correlation network of individual lipid modules generated by WCNA. (C) Dendrogram of lipid modules identified by WCNA. (D) Heatmap showing module associations with AD, MCI, and APOE-ε4 genotype, including AD associations within sex-stratified subsets. Colors depict coefficients from linear regressions adjusted for confounders, unless the confounders were used as predictor variables (age, sampling site, sex, AD status, and APOE-ε4 genotype). (E-H) Percentwise lipid composition for module M5, M7 M9, and M11, respectively. *p < 0.05, and **p < 0.01.

Module eigengenes for 4 of the 11 modules were associated with AD (p < 0.05) after adjustment for sex and APOE-ε4 (with age and sampling site adjusted for residuals) (Figure 1.D and Supplementary table 2). No association was found between module eigengenes and MCI or APOE4 genotype. When participants were stratified by sex, three modules (M5, M7, and M11) were found associated with AD in the female subset while only one module (M9) was associated with AD in the male subset.

Interestingly, two out of the four AD-associated modules (M9 and M11) were mainly consisted of PCs and PEs (Figure 1G-H). Of note, M9 was highly associated with AD in the female subset (p < 0.01) but not in the male subset, whereas M11 was associated with AD in the male subset but not in the female subset. The remaining AD-associated-modules included M7, a module entirely consisted of lysophosphatidylcholines (LPC), and M5, a module comprised of TGs with a few PCs and PEs (Figure 1E-F). The lipids compositions of all modules are provided in Supplementary Figure 1.

Sensitivity analysis showed that M5, M9, and M11 were associated with AD regardless of any adjustments or with any individual potential confounder (Supplementary table 3). On the other hand, M7 only showed association with AD when adjusted for the APOE4 genotype. The sensitivity analysis also included adjustments for total TG levels, however, no differences were observed after adjusting for total TGs.

### 3.3 Lipid association to AD is strongest in women

We selected AD-associated modules (M5, M7, M9 and M11) for further analysis, primarily focusing on the 47 lipids within these modules. Regression analysis revealed that 16 of the 47 lipids were significantly associated with AD after adjusting for age, sex, APOE4 genotype, sampling site, and multiple testing (FDR-adjusted, Figure 2). Interestingly, these findings were not observed in the male-only subset, whereas no lipids were found associated with AD, including the lipids from M9. However, in the female-only subset, 24 of the 47 lipids were associated with AD, including all 16 lipids identified in the full non-subsetted cohort, except for phosphatidic acid (PA)(34:2), which was only associated with AD in the full cohort. No associations were found between these 47 lipids and MCI, MMSE score, or APOE genotype, in neither in the full cohort or in the sex-stratified subset (Supplementary table 4). Of note, most of the selected lipids (37 out of 47) exhibited a strong association with sex after adjustments for other confounders (APOE genotype, age, sampling site and AD status).

**Figure 2.**
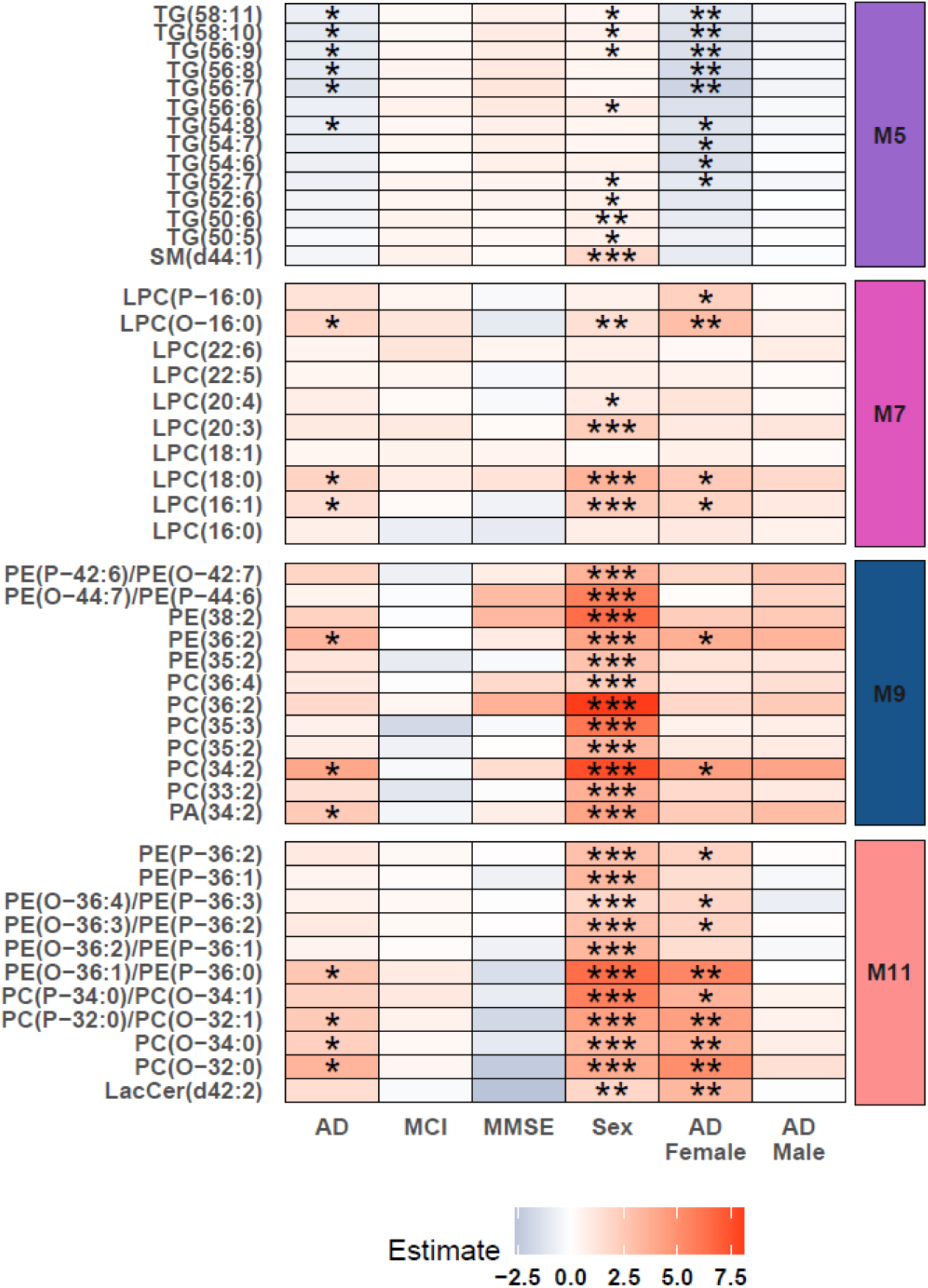
Regression analysis of individual lipids. Heatmap of individual lipid association with AD, MCI, MMSE, APOE-ε4 genotype, sex, and AD in sex-stratified subsets. Colors depict coefficients of linear regressions adjusted for confounders, unless the confounders were used as predictor variable (age, sampling site, sex, AD status, APOE-ε4 genotype). *p < 0.05, **p < 0.01 and *** p < 0.001. P-values are adjusted for multiple testing with FDR.

### 3.4 Lipid association to AD is dependent on degree of unsaturation

We next sought to investigate which lipids are associated with AD. We found a reduction in larger unsaturated lipids, specifically PCs, PEs and TGs, were associated with AD. On the contrary, there was an increase in saturated and mono-unsaturated lipids, particularly from the PC and PE families, which were also associated with AD (Figure 3). We performed a linear regression analysis to understand the association of lipid and AD. After adjusting for sampling site and multiple testing (FDR), we revealed that 37 lipids were associated with AD in the female subset, while no lipids associated with AD in the male subset (Figure 3A). Of these 37 lipids, 14 lipids were positively associated with AD, which included saturated or monounsaturated PCs and PEs, many of which were of the ether variety (-O or -P). In contrast, 23 lipids were negatively associated with AD, they primarily were highly unsaturated TGs, PCs, and PEs. In addition, these lipids showed a stepwise decrease from the controls to MCI, with a larger decrease from the controls to AD, however, this pattern was absent in male participants (Figure 3 B-D). Overall, our data show that there was a tendency for more unsaturated lipids to be negatively associated with AD in female but not in male participants. This trend was particularly pronounced for TGs (Figure 3E). A causal mediation analysis of the 37 lipids associated with AD in women revealed partial mediation. Of these 37 lipids were 15 unsaturated lipids partially mediated by cholesterol (total), LDL, and ApoB (ACME p-value > 0.05). Proportion of mediation ranged from 9 to 66%.

**Figure 3.**
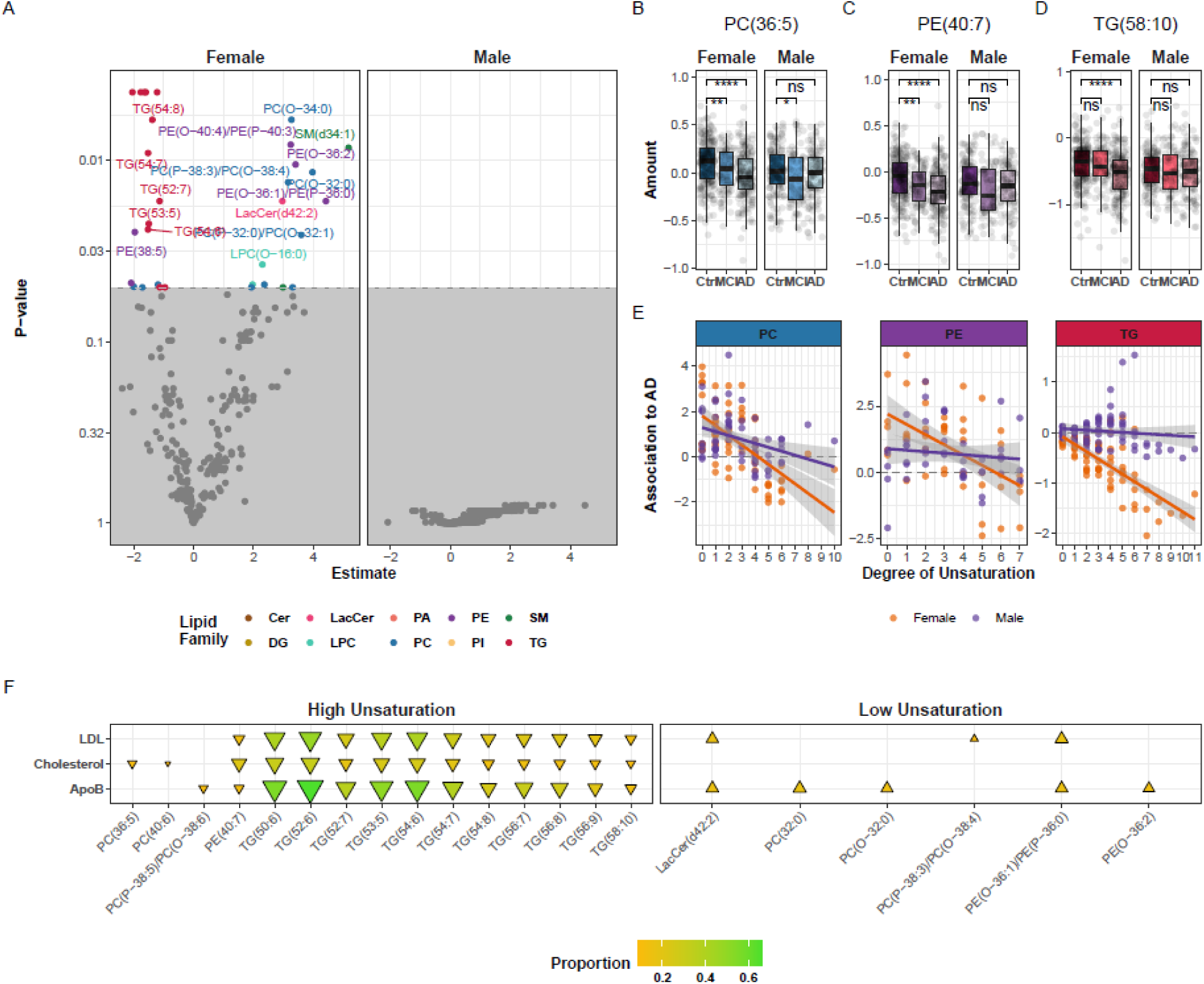
A sex- and saturation-dependent association of lipids with AD. (A) Volcano plots showing lipids differences between participants with AD and cognitively healthy controls, stratified by sex (Left: female subset; Right: male subset). Lipid associations was identified using linear regression adjusted for sampling site and age, with FDR correction for multiple testing. (B-D) Boxplots of selected lipids by cognitive status (Ctrl = cognitively healthy, MCI = mild cognitive impairment, and AD = Alzheimer’s disease) and sex. *p < 0.05, **p < 0.01, ***p < 0.001, and ****p < 0.0001. (E) Unsaturation plot, shows each measured lipid within a lipid family PC, PE or TG, visualizing degree of unsaturation as number of carbon-carbon double bonds and association to AD as the estimate of the linear regression adjusted for sampling site and age. (F) Causal mediation analysis of lipids associated with AD through LDL, total cholesterol, and ApoB. Only lipids with significant associations to AD, the mediator, and an ACME p-value < 0.05 are displayed. The proportion of mediation is represented by the color and size of the triangles. ▲ indicates lipids increased in AD, while ▾ indicates lipids decreased.

In contrast, 6 out of 37 of lipids with low levels of unsaturation, were mediated by LDL and ApoB (Figure 3F). No significant effects were observed between total AD and total TG or HDL, precluding mediation analysis; estimates are provided in Supplementary Table 5.

### 3.4 Lipid difference in male and female participants

We then examined the differences in lipid levels between female and male participants using students *t*-tests, revealing substantial variations in lipidomes between female and male individuals. Overall, lipids levels were generally higher in the female subset, with the levels of SMs particularly elevated in female subset, whereas PE(36:0) was the only lipid found to be higher in male subset (Figure 4A). Among the healthy controls, 179 lipids showed differences between female and male participants after FDR adjustment, while 157 lipids exhibited differences between the sexes in participants with AD. By comparing healthy controls and AD participants, we found that 139 lipids overlapped showing differences, between male and female participants, (Figure 4B). Only 18 lipids showed sex-specific differences exclusively in participants with AD, primarily comprising PC-O, PC-P, PE-O, PE-P, and LPCs (Figure 4C).

**Figure 4.**
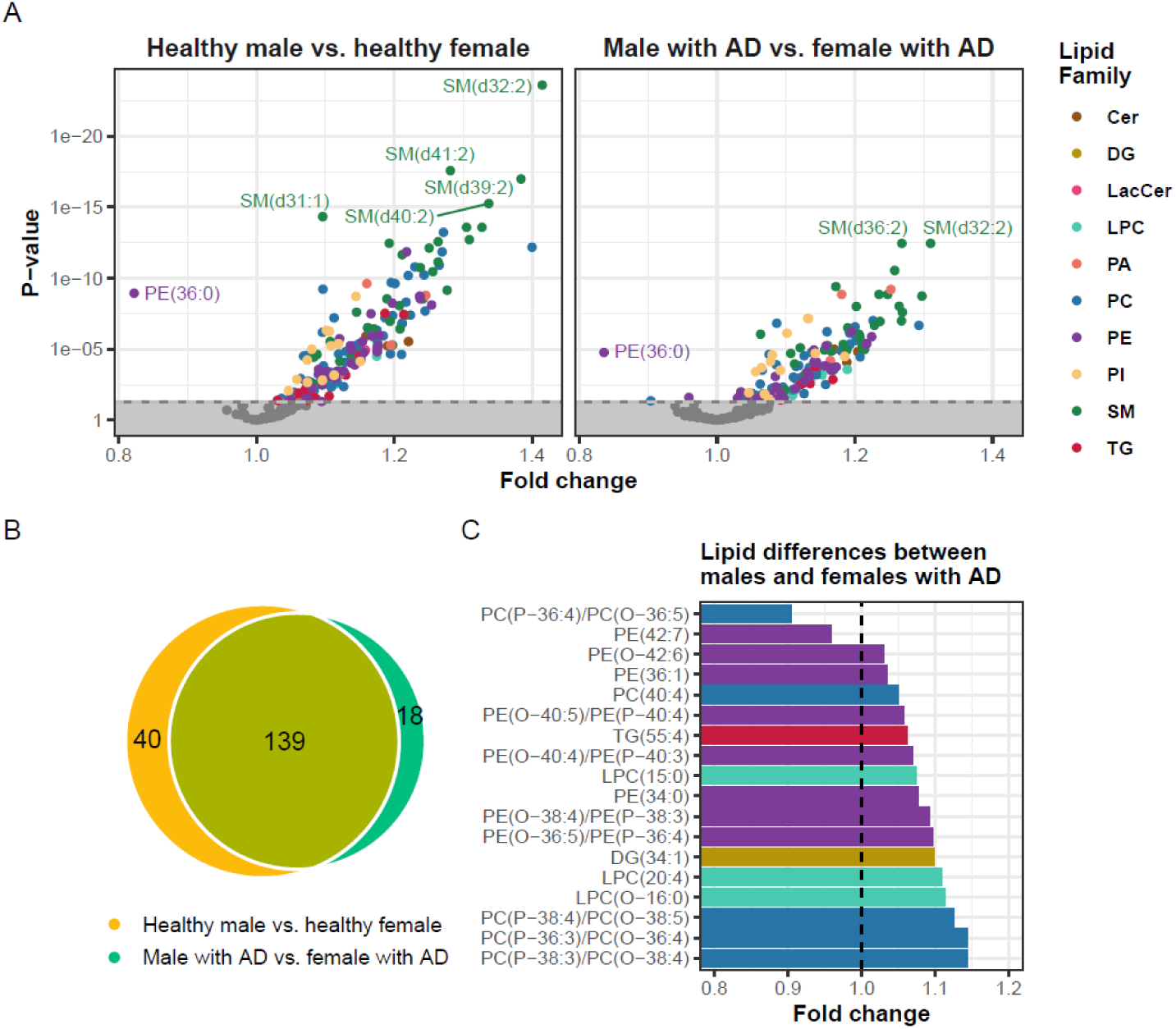
Sex-based lipid differences. (A) Volcano plots showing lipid differences between male and female participants, stratified by AD status (Left: cognitively healthy; Right: AD). Group comparison was performed using students *t*-test, with FDR correction for multiple testing. A positive fold change indicates higher lipid levels in female participants, whereas a negative fold change indicates a higher lipid levels in male participants. (B) Venn diagram of lipids showing the numbers of significantly different lipids between male and female participants, comparing cognitively healthy and AD participants, and lipid different in both groups. (C) Bar plot showing the 18 lipids that differ only between male and female participants with AD and their corresponding fold changes.

## 4. Discussion

This study employed mass spectrometry-based lipidomic analysis and identified 10 lipid families and 268 molecular lipids in the plasma of 841 participants categorized as cognitively healthy, with MCI, or with AD. Our findings reveal significant sex-specific differences in lipid associations with AD, contributing to the growing evidence that AD may manifest differently on the molecular level between women and men. The main difference in women was a deficit of molecular lipids containing polyunsaturated fatty acids, which in turn showed to be partially mediated by cholesterol, LDL and ApoB.

### 4.1 Lipids in women depleted in AD

Our data revealed that, lipid associations with AD were predominantly driven by female participants. Of the 11 lipid modules identified through correlation-based grouping, four were associated with AD in the full cohort. Interestingly, three of these modules showed significant associations in the female subset, while only one was found associated with AD in the male subset. This pattern was more pronounced at the individual lipid level, where 37 lipids were associated with AD in women, while no individual lipids associations were detected in men.

These findings align with those of Lim *et al.* (35) that reported stronger AD-associated lipid changes in women compared to men. Arnold *et al.* (9) observed similar sex-dependent differences in PCs, valine, glycine, and proline, noting these were particularly pronounced in women carrying the APOE4 allele. They proposed that impaired mitochondrial energy production in women with AD might contribute to altered lipid levels.

### 4.2 Decreased levels of highly unsaturated lipids in women with AD

We observed that lower levels of several TGs in module M5 were associated with AD, particularly in women. This observation contrasts with previous studies by Liu *et al.* (12) and Liu *et al.* (36), which found no significant TG changes related to AD or MCI. This discrepancy may be attributed to use of mass spectrometry platform in this study, which enables for more granular analysis of individual lipid species compared to the NMR-based or clinical TG measurements used in previously mentioned studies.

Our study revealed that highly unsaturated lipids, with five or more carbon-carbon double bonds, were reduced in women with AD, primarily TGs, PC and PE. On the other hand, were PCs and PEs with low unsaturation increased in women with AD. A general trend was observed where higher degree of unsaturation had a stronger negative association to AD in women but not in men. The association between highly unsaturated lipids and AD may be linked to the amount of omega-3 fatty acids, such as eicosapentaenoic acid (EPA) and docosahexaenoic acid (DHA), incorporated into the tail ends of these lipids. Notably, women have higher levels of omega-3 fatty acids than men (37). Our findings align with the previous work of Lim *et al.* (35), which reported that cognitively healthy women had higher levels of DHA and omega-3 esterified lipids compared to men, suggesting a link between omega-3 fatty acids and AD. However, this sex-specific difference is absent in AD patients, as shown by a reduction in omega-3 esterified lipids to similar levels in both sexes. Importantly, Lim *et al.* noted that the association between lipids and AD was stronger in women, further highlighting the sex-specific nature of lipid alterations in AD pathogenesis. It should also be noted that the APOE4 genotype influences the amount of DHA in relation to brain volume (38).

Causal mediation analysis of lipids associated with AD revealed that the effects of several highly unsaturated lipids were partially mediated by cholesterol, LDL, and ApoB. These findings align with evidence supporting the potential repurposing of statins to slow cognitive decline in AD (39). Interestingly, the therapeutic effect of statins appears to be greater in individuals carrying the APOE4 allele, consistent with findings from Fu *et al*., which show that cholesterol and LDL levels are higher in APOE4 carriers, with this difference being more pronounced in women (40). In this study we found no lipids associated with APOE4 carriers.

Another avenue to explore is hormone replacement therapy, which have showed cognition improvement in women carrying APOE4 variant (41), it would be interesting to investigate whether these improvements also was reflected in the levels of highly unsaturated lipid, DHA or cholesterol.

### 4.3 Increases in PC-O and PC-P (plasmalogen’s) and their anti-inflammatory roles

Higher levels of individual low unsaturation PCs and PEs lipids were associated with AD in women but not in men. These findings align with previous studies by Lim *et al.* (35), which found an increased level of some low unsaturation PCs and PEs species associated with AD and a decreased level of higher unsaturation PC and PEs associated with AD (35), a pattern also observed and Whiley *et al*. (22).

Two of the four AD-associated modules (M9 and M11) primarily consisted of PCs and PEs. We revealed that M9 was associated with AD in men but not in women, while M11 showed the opposite pattern. M11 was unique in that it exclusively contained ether-carrying PCs and PEs: alkyl ether substituent (-O) and 1Z-alkenyl ether substituent (-P), also known as plasmalogens. Plasmalogens have been noted for their anti-inflammatory effects especially in neurodegeneration (42–44). They are depleted with age (45) and in the brains of AD patients (46,47). Our finding of increased plasmalogen levels in blood could be a sign of an anti-inflammatory response to AD. The role and mechanism of plasmalogens in relation to AD warrants further investigation.

### 4.4 Increase in LPCs

In our network analysis, we found that LPCs showed minimal correlation with other lipid families, which results in a distinct module solely comprised of LPCs, that was associated with AD in both the full cohort and the female subset but not in the men subset, Klavins *et al*. also observed an nominal increase of LPC species in people with AD (48). LPCs deliver unsaturated lipids, such as DHA, into the brain by utilizing the sodium-dependent LPC symporter 1 to transport them across the blood-brain barrier (49), higher levels of LPCs in the blood could be a consequence of lower amount of unsaturated fatty acids to transport. This finding highlights the importance of LPCs in AD pathogenesis, particularly in women.

One limitation is that this cohort consists of elderly participants primarily of European descent, with selection criteria that may favor a specific AD pathology. Larger, multi-ethnic validation studies are needed to be able to generalize these results. Another limitation of our study is that the specific position of carbon-carbon double bond could not be determined, making it is challenging to differentiate between certain lipid types, such as distinguishing a PC-O with one additional carbon-carbon double bond and a PC-P

In conclusion, our study demonstrates that associations between various lipid species (TGs, PCs, PEs, and LPCs) and AD are more prominent in women and often absent in men. These findings highlight the importance of sex-stratified analyses in AD research, which could contribute to the identification of sex-specific molecular mechanisms. Future studies should further investigate the mechanistic underpinnings of these sex-specific lipid alterations and their potential for targeted intervention in individuals with AD.

## Abbreviations

AD: Alzheimer’s disease
ADE: Average direct effect
ACME: Average causal mediation effect
Cer: Ceramides
CDR: Clinical Dementia Rating Scale
Ctrl: Control
DHA: Docosahexaenoic acid
EDTA: Ethylenediamine tetraacetic acid
FDR: False discovery rate
HDL: High density lipoprotein
KNN: K-nearest neighbor
LDL: Low density lipoprotein
LPC: Lysophosphatidylcholines
MTBE: Methyl tert-butyl ether
MCI: Mild cognitive impairment
MMSE: Mini-Mental State Examination
M1-12: Module 1-12
NINCDS-ADRDA: National Institute of Neurological and Communicative Disorders and Stroke and the Alzheimer’s Disease and Related Disorders Association
PA: Phosphatidic acid
PC: Phosphtatidylcholines
PE: Phosphtatidylethanolamines
QTOF-MS: Quadrupole time-of-flight mass spectrometer
SM: Sphingomyelin
TG: Triglycerides
UPLC: Ultra-performance liquid chromatography
WCNA: Weighted correlation network analysis

## Statements

### Author Contribution

Following the Contributor Role Taxonomy (CRediT). **A.W.:** Conceptualization, Formal analysis, Methodology, Project administration, Software, Visualization, Writing - original draft, Writing - review & editing. **J.X.:** Data curation, Investigation, Writing - review & editing. **W.C.:** Writing - review & editing. **L.V.:** Data curation, Resources, Writing - review & editing. **P.P.:** Conceptualization, Funding acquisition, Project administration, Writing – review & editing. **C.L.-Q.:** Conceptualization, Funding acquisition, Project administration, Supervision, Writing – review & editing.

### Ethical approval

Informed consent was obtained for all subjects according to the Declaration of Helsinki (1991), and protocols and procedures were approved by the relevant local ethical committees.

### Availability of data and materials

The dataset analyzed in this study contains sensitive participant information and is not publicly available due to privacy concerns. However, qualified researchers may request access to the data through the AddNeuroMed consortium. All code used for data analysis is openly accessible via GitHub: https://github.com/Asger-W/AddNeuroMed-Lipidomics.

