## Supplementary Figures for "Lipid profiling reveals a deficit of lipids with unsaturated fatty acids in women with Alzheimer’s disease"

### Supplementary material

Supplementary Tables

The following tables can be found in the accompanying spreadsheet:

Sup. Table 1 – Participant characteristics by sex

Sup. Table 2 – Module Eigengene Regression

Sup. Table 3 – Module Eigengene AD Sensitivity Analysis

Sup. Table 4 – Individual lipid Regression

Sup. Table 5 – Causal mediation analysis

Supplementary Figures

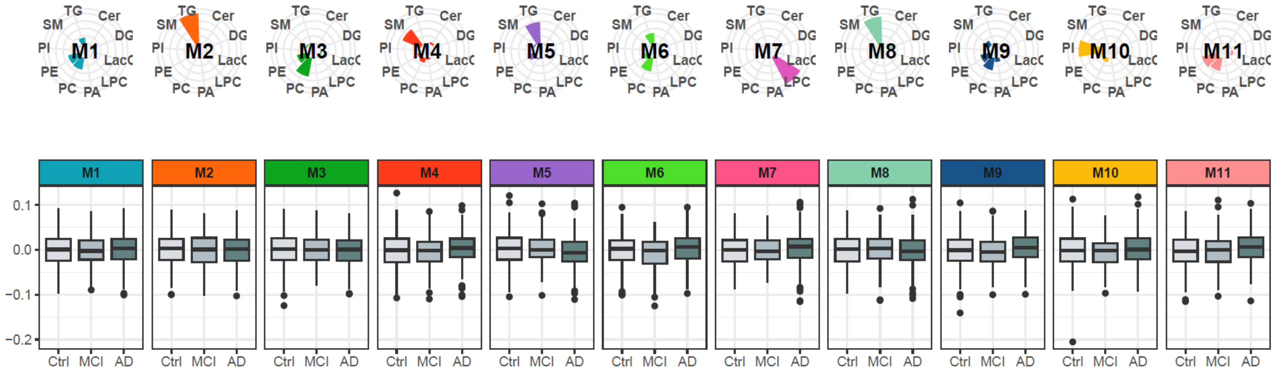


Sup. Figure 1. Full module composition and Eigengene boxplots by AD status and Sex. Percentwise lipid composition of each module, along with eigengene boxplot by cognition status (Ctrl = cognitively healthy, MCI = mild cognitive impairment, and AD = Alzheimer's disease).
